# Primary Amoebic Meningoencephalitis caused by Complement C2 Deficiency

**DOI:** 10.64898/2026.01.31.26345168

**Authors:** Jian Cui, Colleen M. Roark, Nerea Domínguez-Pinilla, Pilar Nozal Aranda, Begoña Losada, Pilar Zamarrón, Jacob Lorenzo-Morales, José Miguel Rubio Muñoz, Megan M. Dobrose, Ana Van den Rym, Luis M. Allende, Catherine Shelton, Dante E. Reyna, Janet G. Markle, Santiago Rodríguez de Córdoba, Margarita Lopez-Trascasa, Rebeca Pérez de Diego, C. Henrique Serezani, Mariana X Byndloss, Isabel de Fuentes Corripio, Luis Ignacio González-Granado, Ruben Martinez-Barricarte

**Affiliations:** Division of Genetic Medicine and Clinical Pharmacology, Department of Medicine, Vanderbilt University Medical Center, Nashville, TN, USA; Division of Molecular Pathogenesis, Department of Pathology, Microbiology and Immunology, Vanderbilt University Medical Center, Nashville, TN, USA; Pediatric Hematology and Oncology Unit. University Hospital 12 de Octubre. Research Institute Hospital 12 Octubre (imas12). Madrid, Spain; Department of Immunology, La Paz University Hospital, Madrid, Spain; Complement Alterations in Human Pathology Group, La Paz Institute of Biomedical Research, Madrid, Spain; Pediatric Unit, University Hospital of Toledo, Spain; Microbiology service, Hospital Virgen de la Salud, Toledo, Spain; University Institute of Tropical Diseases and Public Health of the Canary Islands, University of La Laguna, La Laguna, Spain; Department of Pediatric Obstetrics and Gynecology, Pediatrics, Preventive Medicine and Public Health, Toxicology, Legal Medicine, Forensics and Parasitology, University of La Laguna, La Laguna, Spain; CIBER on Infectious Diseases (CIBERINFEC), Carlos III Health Institute, Madrid, Spain; Malaria & Emerging Parasitic Diseases Laboratory, Parasitology Department, National Microbiology Centre, Carlos III Health Institute, Majadahonda, Spain; Laboratory of Immunogenetics of Human Diseases, IdiPAZ Institute for Health Research, La Paz University Hospital, Madrid, Spain; Department of Immunology, University Hospital 12 de Octubre, Madrid, Spain; Research Institute Hospital 12 Octubre (imas12), Madrid, Spain; School of Medicine, Complutense University of Madrid, Madrid, Spain; Vanderbilt Institute of Infection, Immunology and Inflammation, Vanderbilt University Medical Center, Nashville, TN, USA; Division of Infectious Diseases, Department of Medicine, Vanderbilt University Medical Center, Nashville, TN, United States; Vanderbilt Center for Immunobiology, Vanderbilt University Medical Center, Nashville, TN, USA; Vanderbilt Genetics Institute, Vanderbilt University Medical Center, Nashville, TN, USA; Genetic and Molecular Complement Diagnostic Laboratory, Center for Biological Research Margarita Salas, Madrid, Spain; Department of Medicine, Autonoma University of Madrid, Madrid, Spain; Howard Hughes Medical Institute, Vanderbilt University Medical Center, Nashville, TN, USA; Toxoplasmosis and intestinal protozoan unit, Referral and investigation lab in parasitology, National Center of Microbiology, Carlos III Health Institute. Majadahonda, Spain; Department of Pediatrics, University Hospital 12 de Octubre, Research Institute Hospital 12 Octubre (imas12). Madrid, Spain

**Keywords:** Primary amoebic meningoencephalitis (PAM), *Naegleria fowleri*, innate immunity, complement system, complement C2 deficiency, Inborn error of immunity (IEI)

## Abstract

**Background:** Primary amoebic meningoencephalitis (PAM) is a rapidly progressive and often fatal central nervous system infection caused by *Naegleria fowleri*. Despite widespread environmental exposure to this free-living amoeba, clinical disease is rare, suggesting that it requires not only exposure to the amoeba but also a host vulnerability. Yet, the immune mechanisms controlling protection vs. susceptibility to *N. fowleri* remain poorly understood.

**Methods:** We conducted comprehensive clinical, immunological, and genetic investigations in one of the few survivors of PAM. We performed high-dimensional immune profiling using Cytometry by Time-Of-Flight (CyTOF) to assess immune cell composition and activation state. We employed whole-exome sequencing (WES) to identify rare genetic variants that affect host responses. Functional immune assays were used to assess serum-mediated amoebicidal activity *in vitro* and to characterize key host defense pathways.

**Results:** A previously healthy pediatric patient was diagnosed with PAM. Contrary to other cases, her clinical course lasted for more than 2 months before she recovered with miltefosine treatment. Immunologic evaluation showed this patient had normal numbers and frequencies of major lymphoid and myeloid immune cells. WES revealed a homozygous deletion in the complement component 2 (C2) gene, resulting in a complete absence of circulating C2 protein and abolishing classical complement pathway activity. Normal human serum induced complement-mediated lysis of *N. fowleri* trophozoites in vitro, whereas complement-depleted normal human serum and serum from our patient both failed to deposit membrane attack complex (MAC) or kill *N. fowleri*. MAC deposition and amoebicidal activity were restored by supplementing the patient’s serum with purified human C2 protein.

**Conclusion:** Our study demonstrates that PAM can be caused by a monogenic inborn error of immunity (IEI) and that the complement system is critical for human immunity against *Naegleria fowleri*.

## Introduction

Primary amoebic meningoencephalitis (PAM) is a rare but almost universally fatal infection of the central nervous system caused by *Naegleria fowleri* (*N. fowleri*), a thermophilic, free-living amoeba commonly found in warm freshwater environments such as lakes, rivers, hot springs, and untreated swimming pools worldwide.^1–4^ Rising global temperatures have expanded the geographic range of *N. fowleri*, increasing potential exposure in previously unaffected regions.^5–7^ *N. fowleri* infections are commonly linked to recreational activities like swimming or diving, but can occur in any situation in which contaminated water comes into contact with the host’s nasal epithelium.^8–11^ The amoeba can exist as dormant cysts, a transient flagellate stage, and infectious trophozoites. (Figure 1A). In a minority of exposed individuals, once in contact with the nasal epithelium, trophozoites migrate along the olfactory nerve to the brain, where they can proliferate and induce an acute inflammatory reaction, leading to hemorrhagic and necrotizing meningoencephalitis.^12,13^ (Figure 1A)

**Figure 1:**
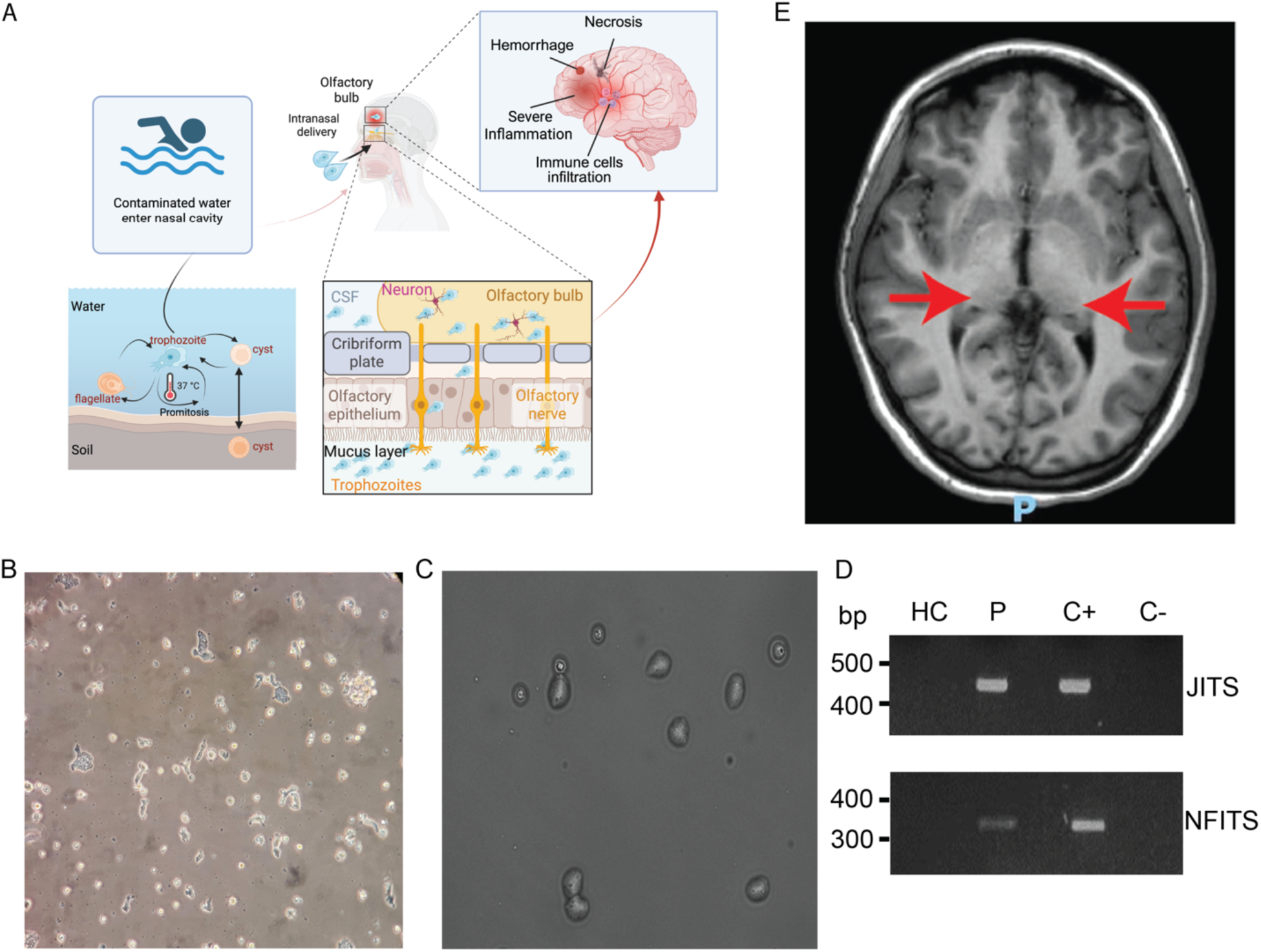
*N. fowleri* life cycle and PAM diagnosis in a patient. (A) *N. fowleri* life cycle and pathogenesis. Free-living trophozoites in warm freshwater penetrate the nasal mucosa during water exposure and migrate through the olfactory epithelium. The organism travels through the cribriform plate, to the olfactory bulbs and cerebrospinal fluid (CSF), leading to central nervous system invasion. B) Trophozoite and (C) cyst observed in *N. fowleri* cultured from the patient’s CSF. (D) PCR amplification of *N. fowleri* ITS in CFS from a Healthy Control (HC), patient (P), *N. folweri* positive control (C+), and negative control (C-). JITS primers are used in the upper panel and NFITS primers in the lower one. (E) Magnetic Resonance Imaging (MRI) of the patient. Small porencephalic lesions located in both thalami, slightly larger on the right side, where they measure approximately 15 mm, without contrast enhancement. Focal lesion, without mass effect, in the left hemipons (less than 1 cm in size, with no contrast enhancement).

Clinically, PAM presents with sudden fever, headache, nausea, vomiting, and altered mental status, often progressing swiftly to coma and death within 3 to 7 days after the onset of symptoms.^14–16^ The infection’s rapid and fatal course severely restricts opportunities to study host defense during active disease, and the rarity of survivors (11 reported globally to date) further limits the potential to investigate protective immune responses. Diagnosis is typically delayed and relies on nonspecific clinical symptoms or microscopic detection of amoebae in cerebrospinal fluid — a method that is neither sensitive nor timely - while treatment is similarly non-targeted, relying on empiric combinations of antimicrobial and antifungal agents with unproven and largely inconsistent benefit.^3,9,10^ Autopsy studies have revealed extensive inflammatory destruction of brain tissues but provided little information on early immune events or mechanisms that might contribute to survival.^17,18^ Identifying host factors that determine susceptibility vs. protection is critical to improving diagnosis, prevention, and treatment.

Despite evidence of widespread exposure, with seropositivity exceeding 90%, clinical disease caused by N. fowleri remains exceedingly rare, suggesting that host immune defenses prevent infection in the vast majority of exposed individuals.^19–22^ The apparent rarity and selective susceptibility to PAM in previously healthy children and young adults^23,24^, raises the possibility that PAM may be an unrecognized inborn error of immunity (IEI). IEIs comprise a heterogeneous group of monogenic disorders that result in selective or broad susceptibility to infections, autoimmunity, or malignancy.^25,26^ Although IEIs have been increasingly recognized as underpinning severe infection by individual pathogens, most severe infectious diseases have not yet been investigated as potential IEI. Here, we describe a documented survivor of PAM beyond the 11 previously reported cases. By studying this patient through the lens of IEI, we discover that susceptibility to *N. fowleri* can be a monogenic disease and identify critical and non-redundant players in human anti-*N. folweri* immunity.

## Materials and Methods

### Cytometry by Time-of-Flight (CyTOF)

Human PBMCs were isolated by Ficoll-Hypaque density gradient centrifugation (Amersham-Pharmacia-Biotech, Buckinghamshire, UK) from whole-blood samples obtained from the patient, parents, and healthy donors, and cryopreserved in FBS containing 10% DMSO (VWR, Radnor, PA, USA). 3 million peripheral blood mononuclear cells (PBMCs) were stained with Cell-ID Cisplatin-195Pt (Standard BioTools, South San Francisco, CA, USA) to discriminate dead cells. Human TruStain FcX (Biolegend, San Diego, CA, USA) was used to block Fc receptors, and PBMCs were stained with the antibody mix as shown in Table S1. Cells were fixed with 1.6% paraformaldehyde and stained with Cell-ID intercalator-Ir. Stained PBMCs were analyzed on a Helios Mass Cytometer at the University of Maryland Mass Cytometry Core Laboratory. The data were exported as a Flow Cytometry Standard (FCS) file and normalized using EQ bead standards according to the manufacturer’s protocol. Data analysis was performed as described previously. ^27^

### Genetic Analysis

Genomic DNA (gDNA) was isolated from the whole blood of the patient and his parents using the GeneJET Genomic DNA Purification Kit (Thermo Fisher Scientific, Waltham, MA, USA). Whole-exome sequencing (WES) was performed on an Illumina NovaSeq 6000 platform. Briefly, library preparation and capture were performed utilizing the XGen Research Panel probes from IDT according to the manufacturer’s instructions. Paired-end sequencing (2 × 150 bp) was conducted, targeting an average of 20 million reads per sample (∼50× coverage). Bioinformatic analysis was performed using Webseq as previously described.^28^ Primers were designed to amplify exon 5 and intron 6 of *C2* containing the mutation of interest (Supplementary Table S2). A PCR was set up with these primers and gDNA from the patient, family, and a healthy donor using DreamTaq Green PCR Master Mix (2X) (Thermo Fisher Scientific). Reactions were incubated in a thermal cycler according to product specifications and run on a 1% agarose gel to confirm amplification. PCR products were Sanger sequenced (Genewiz Azenta Life Sciences, South Plainfield, NJ, USA) using the primers shown in Supplementary Table S2.

### Complement Component Quantification

Serum concentrations of C1q, C2, C3, C4, C5, MBL, Factor B, Properdin, Factor H, and Factor I were quantified using the Human Complement 15-Plex Discovery Assay® (Eve Technologies, Calgary, AB, Canada).

### Complement Pathway Functional Assays

Classical and alternative pathway activities were measured using standard hemolytic assays based on the lysis of antibody-sensitized sheep erythrocytes (CH50) or unsensitized rabbit erythrocytes (AH50) (Complement Technology, Inc., Tyler, Texas, USA) following the manufacturer’s instructions. C2-deficient serum was reconstituted with increasing concentrations of purified human C2 protein (Complement Technology, Inc.), followed by a CH50 assay to evaluate restoration of classical pathway activity.

### Naegleria fowleri Culture

*Naegleria fowleri* Carter (ATCC 30894, Manassas, Virginia, USA) trophozoites, a pathogenic strain originally isolated from a human case of primary amoebic meningoencephalitis,^29^ were maintained axenically in Nelson’s medium supplemented with 10% fetal bovine serum at 37°C in 75 cm² tissue culture flasks.^30^ Cultures were kept in logarithmic growth phase and counted using a hemocytometer before use in the experiment.

### *Naegleria folweri* identification in CSF by PCR

PCR on CSF from the patient and appropriate controls was performed using specific primers (Supplementary Table S3) to amplify the Internal Transcribed Spacers (ITS), followed by PCR with primers specific to *Vahlkamphfiidae* (JITS) and to *N. fowleri* as described before.^31^

### Complement-Dependent Cytotoxicity Assay

100μl of trophozoites at a concentration of 1 million/ml were seeded in a 96-well cell culture plate. After overnight incubation, trophozoites were incubated with 30% human serum from the patient, healthy controls, unrelated C2-deficient individuals, or commercial C2-depleted serum (Complement Technology, Inc.). To assess C2-dependent phenotypes, sera were pre-treated with the factor B inhibitors Iptacopan (MedChemExpress, Monmouth Junction, NJ, USA) or FB28^32^ for 1 hour at room temperature to partially block alternative pathway activation, which would otherwise mask any classical or lectin pathway contribution to amoebicidal activity. Heat-inactivated (56°C for 30 minutes) and C5-depleted serum (Complement Technology, Inc.) served as the negative control. For reconstitution, purified human C2 protein was added to factor B inhibitor-treated patient serum. After 30 minutes of incubation at 37°C, cell death was assessed by propidium iodide (PI) staining (Thermo Fisher Scientific). MAC deposition was evaluated by flow cytometry and fluorescence microscopy using anti-C9 (E-3) Alexa Fluor® 488 (Santa Cruz Biotechnology Inc., Dallas, Texas, USA).

### Flow Cytometry

Following serum incubation as mentioned above, trophozoites were washed and stained with PI and anti-C9 (E-3) Alexa Fluor® 488. Data were acquired using a BD LSR Fortessa cytometer (BD Biosciences, San Jose, CA, USA) and analyzed with FlowJo software (BD Biosciences).

### Immunofluorescence Microscopy

DAPI (Thermo Fisher Scientific), DRAQ7 (Thermo Fisher Scientific), and anti-C9 (E-3) Alexa Fluor® 488 were added to the serum before incubation to assess MAC deposition and nuclear integrity. Images were captured and analyzed using Cytation 7 (Agilent Technologies) with identical acquisition settings across samples to ensure comparability.

### Electron Microscopy

Trophozoites treated with NHS were fixed in 2.5% glutaraldehyde and extensively washed in 0.1 M cacodylate. After fixation, samples were adhered to poly-lysine-treated Si wafers for 15 minutes and sequentially postfixed in 1% tannic acid, 1% OsO4, and 1% uranyl acetate. The wafers were dehydrated in a graded ethanol series and critical point dried in a Tousimis PVT-3D, followed by sputter coating with platinum using a Cressington 108 sputter coater. Naegleria fowleri was imaged using a Zeiss Crossbeam 550 FIB-SEM at 2 keV using the lower secondary electron detector. Tile sets were acquired and stitched using Atlas 5 acquisition software.

### Statistical Analysis

Data were analyzed using GraphPad Prism 10. Comparisons between groups were made using unpaired two-tailed t-tests or one-way ANOVA with Tukey’s post hoc test, as appropriate. Results are reported as mean ± SEM. A p-value < 0.05 was considered statistically significant.

## Results

### Clinical presentation and treatment of a PAM survivor

We present a pediatric female patient with a history of recurrent ear infections during early childhood and an appropriate response to routine vaccinations. She was born to healthy, non-consanguineous parents and has a healthy sister. She presented with progressive left retro-orbital pain, left trapezius muscle pain, and occipital neuralgia, followed by signs of sixth cranial nerve palsy. Her neurological status progressively deteriorated during the following months until *N. fowleri* growth was observed in her CSF and confirmed by PCR (Fig. 1B-D). At this point, she was started on miltefosine and recovered. Follow-up MRI (Figure 1E) demonstrates residual lesions involving both thalami and the left hemipons. She is currently independent in activities of daily living.

### Immunophenotyping revealed normal immune cell counts and frequencies in peripheral blood

Given the severity of PAM, we hypothesized that susceptibility may result from an underlying defect in essential immune cells required for effective host defense against *N. fowleri*. To test this, we performed high-dimensional immunophenotyping using CyTOF on PBMCs from the patient (P1), one of her parents, and healthy controls. By unsupervised clustering, we identified major immune lineages—including CD4⁺ and CD8⁺ T cells, B cells, natural killer (NK) cells, myeloid cells, and dendritic cells—confirmed by canonical surface marker expression (Fig. 2A–B). The patient’s immune cell landscape was similar to that of the healthy control and her parent (Fig. 2A–C). Quantitative analysis revealed normal absolute counts and relative frequencies across 35 major lymphoid and myeloid cell populations (Fig. 2D, S1, Table 1. No aberrant clustering or expansion of atypical populations was detected, suggesting similar immune cell composition between healthy controls and the patient. Overall, these findings indicate that the patient’s peripheral immune system is phenotypically intact, with no evidence of global cellular immunodeficiency.

**Figure 2:**
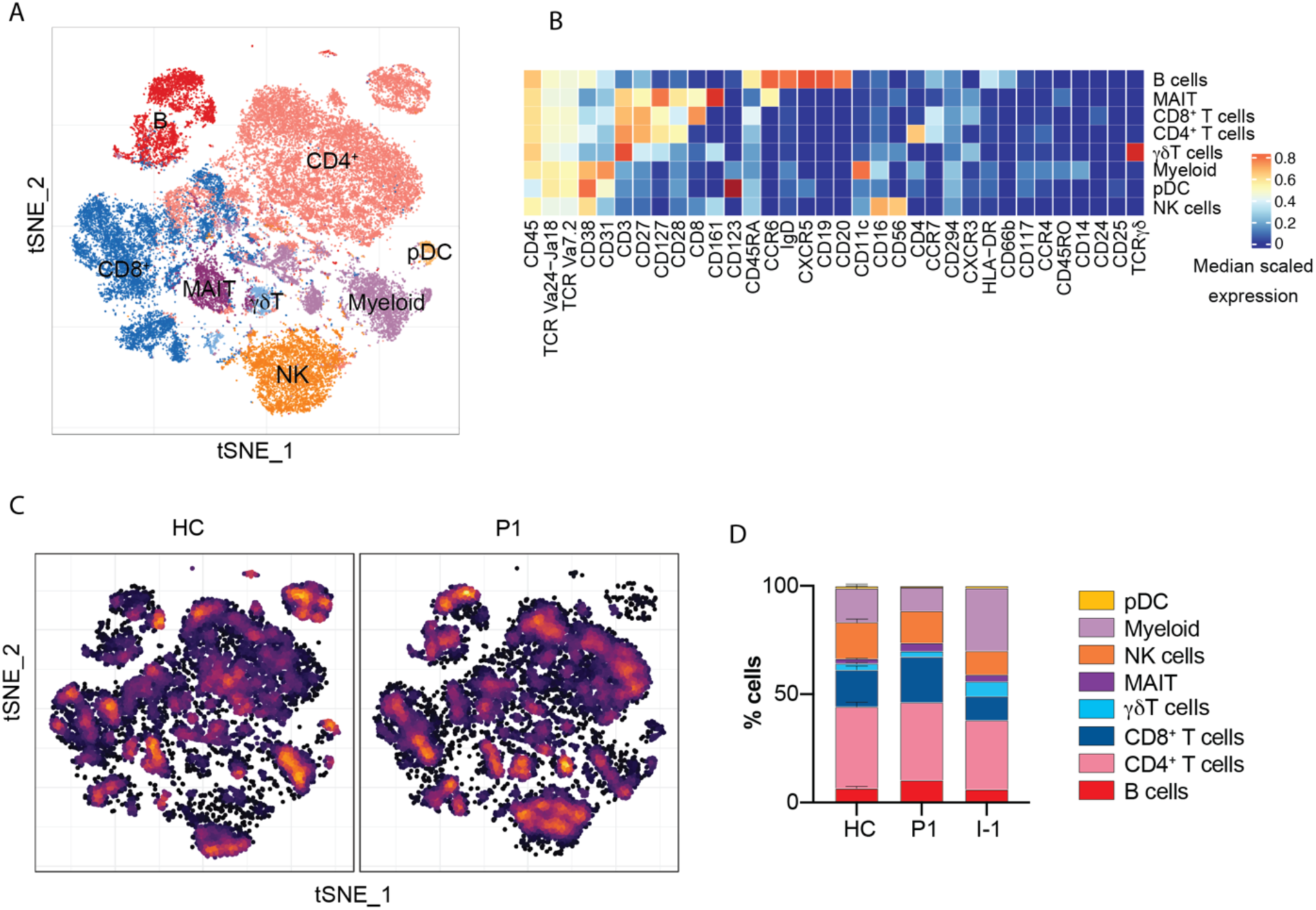
In-depth immunophenotyping of P1 by cytometry by time of flight. (A) Dimensional reduction by t-SNE of the 34 cell surface markers used for mass cytometry immunophenotyping. Each color denotes manually clustered immune cell types. (B) Heatmap showing marker expression of the populations in (A). (C) Density t-SNE comparing cell distribution between the patient and a representative healthy control(A). (D) Immune cell frequencies as a percentage of total leukocytes for healthy controls (HC), the patient (P1), and a patient’s parent (I-1).

**Table 1:** Immune cell frequencies in a patient with PAM.

| **Population (% of leukocytes)** | **HC** | **Patient** |
| --- | --- | --- |
| B cells | 5.26-14.5 | 10.3 |
| Plasmablasts | 0.056-0.3 | 0.14 |
| CD4 T cells | 25-42.6 | 28.2 |
| $\gamma\delta$T cells | 0.46-3.4 | 3.64 |
| Follicular T cells | 0-8.39 | 7.7 |
| Recent thymic emigrants | 0-15.2 | 9.04 |
| CD8 T cells | 12-27.7 | 15.5 |
| Classical monocytes | 0.33-10.1 | 2.06 |
| Nonclassical monocytes | 0.004-1.58 | 0.27 |
| Intermediate monocytes | 0-0.24 | 0.084 |
| NK cells | 4-23.3 | 11.4 |
| Myeloid DC | 1.78-10.1 | 2.39 |
| Plasmacytoid DC | 0.078-0.55 | 0.17 |
| Total ILCs | 0.33-5.73 | 0.6 |
| **Population (% of B cells)** | **HC** | **Patient** |
| Naïve B cells | 28.7-75.9 | 54.4 |
| Unswitched B cells | 4.74-27.5 | 35.5 |
| Switched B cells | 5.15-34.9 | 6.93 |
| DN B cells | 3.93-13.2 | 3.14 |
| **Population (% of CD4)** | **HC** | **Patient** |
| Naïve CD4 T cells | 56.7-79.4 | 64.1 |
| TEMRA CD4 T cells | 3.7-18.6 | 4.85 |
| EM CD4 T cells | 0.19-7.92 | 5.77 |
| CM CD4 T cells | 0.58-17.4 | 32.1 |
| Th1* cells | 0-2.1 | 0.73 |
| Th17 cells | 1.6-7.06 | 2.72 |
| Th1 cells | 0-2.73 | 2.2 |
| Th2 cells | 2.17-7.37 | 5.16 |
| Regulatory T cells | 1.73-3.83 | 2.69 |

| **Population (% of CD8)** | HC | Patient |
| --- | --- | --- |
| Naïve CD8 T cells | 59.9-83.5 | 66.4 |
| TEMRA CD8 T cells | 3.95-15.8 | 7.98 |
| EM CD8 T cells | 0-5.1 | 6.47 |
| CM CD8 T cells | 0-12.9 | 12 |
| MAIT | 0.36-16.8 | 3.41 |

| **Population (% of NK)** | HC | Patient |
| --- | --- | --- |
| CD56<sup>bright</sup> NK cells | 0.38-2.34 | 1.24 |
| CD56<sup>dim</sup>CD16<sup>+</sup> NK cells | 16.7-89.8 | 68.6 |
| CD56<sup>dim</sup>CD16<sup>-</sup> NK cells | 8.31-83 | 29.6 |

### The patient has a homozygous *C2* mutation, resulting in complete C2 deficiency

Considering the rarity of PAM despite widespread environmental exposure to *N. fowleri*,^19–22^ we hypothesized that the patient’s susceptibility might result from an undiagnosed IEI. To investigate this hypothesis, we performed WES on the patient and her parents. We assumed complete penetrance; hence, we focused on rare (MAF<0.01) variants and performed separate analyses to detect *de novo* variants and those inherited in an autosomal recessive manner (Either homozygous or compound heterozygous). The patient was found to be homozygous for a 28-bp deletion in C2 (c.841_849+19del), which encodes complement component 2 (Figure 3A). This deletion spans the end of exon 6 and intron 6 of C2, removing a donor splice site (Figure 3B), which would predictively affect normal splicing and protein expression. It is classified as Pathogenic/Likely pathogenic in ClinVar (https://www.ncbi.nlm.nih.gov/clinvar/) with multiple independent clinical submissions, and functional evidence indicates it causes autosomal recessive C2 deficiency.

**Figure 3.**
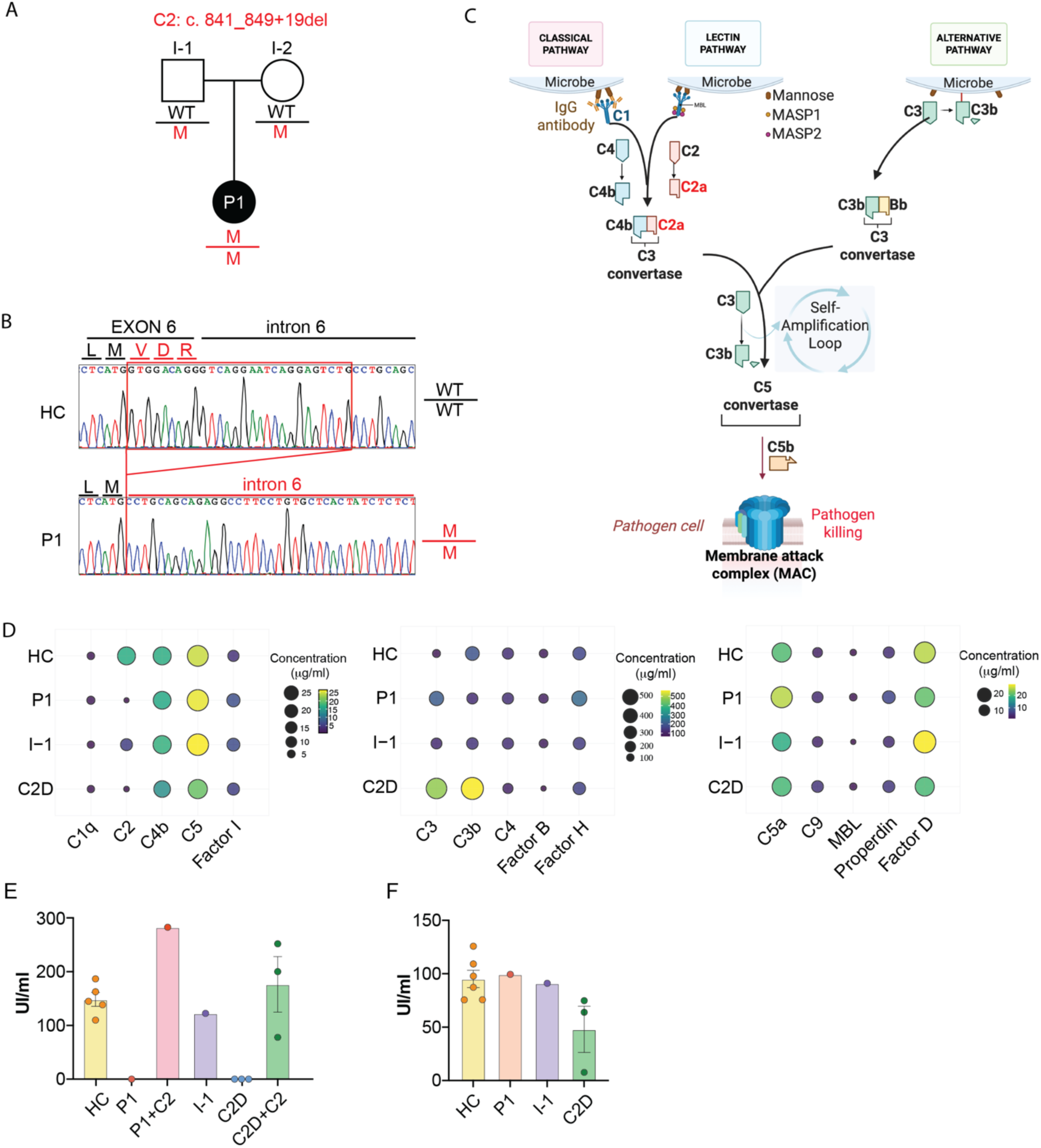
Genetic and Functional Characterization of Complement C2 Deficiency in a PAM patient. (A) Pedigrees of the patient with homozygous C2 deficiency (c.841_849+19del). A solid symbol indicates the affected individual. (B) Sanger sequencing results of the genomic area of *C2* where the mutation is located at the end of exon 6 and start of intron 6 in healthy donors (HC) and the patient (P1). (C) Simplified overview of the complement system. (D) Serum complement components levels in healthy donors (HC), the patient (P1), the patient’s parent (I-1), and unrelated C2-deficient (C2D) patients. (E) Classical pathway activity measured by CH50 assays. (F) Alternative pathway activity measured by AH50 assays. Data represent three independent experiments; mean ± SEM.

The complement system is a crucial part of innate immunity, composed of over 30 plasma and membrane-bound proteins that interact in a tightly regulated proteolytic cascade.^33^ The C2 protein functions in both the classical and lectin pathways, playing a critical role in their activation that culminates in the assembly of the Membrane Attack Complex (MAC) (Figure 3C).^34,35^ We assessed serum C2 protein levels and, consistent with previous reports, found them to be undetectable in the patient’s serum (Figure 3D, S2B).^36,37^ Despite C2 deficiency, upstream classical/lectin initiators, core complement components, alternative pathway factors, and terminal pathway proteins were all within normal ranges, indicating largely preserved complement function (Figure 3D, S2B). Functional complement activity was then assessed using hemolytic and immunoassay-based approaches. The classical pathway activity was absent in the patient’s serum as measured by the CH50 assay, confirming a functional defect in this pathway. Remarkably, the addition of purified C2 restored classical pathway activity (Figure 3E), demonstrating that C2 deficiency was the only driver of the defect. In contrast, the alternative pathway remained fully functional, as demonstrated by a normal AH50 response (Figure 3F). These findings show that our patient has a complete C2 deficiency.

### Complement directly kills *N. fowleri*

To determine whether the complement system plays a critical role in anti-N. fowleri immunity, we evaluate complement-mediated amoebicidal activity using human serum samples. Intact normal human serum (NHS) rapidly and efficiently killed N. fowleri trophozoites, resulting in a greater than 90% reduction in viable trophozoites within 30 minutes of exposure and a robust deposition of C9—the pore-forming component of the MAC—on amoebae (Figure 4A–C). To confirm that the complement system specifically mediated this potent amoebicidal effect, we tested heat-inactivated serum—where complement proteins are denatured—and serum deficient in complement component C5, a key terminal pathway protein required for MAC formation. Both heat-inactivated and C5-deficient sera failed to induce significant C9 deposition or *N. flowleri* killing, with trophozoite survival rates comparable to those of untreated controls (Figure 4A–C). Correspondingly, ultrastructural examination by scanning electron microscopy revealed striking morphological changes exclusively in amoebae treated with NHS, including swelling, loss of amoebastomes, and eventual plasma membrane rupture, consistent with complement-mediated lysis (Figure 4D, S4D). Amoebae treated with heat-inactivated serum or left untreated retained intact membranes and normal ultrastructure (Supplementary figure S4D). These quantitative and morphological data collectively demonstrate that human complement, through formation of the MAC, is a critical innate immune effector that directly mediates rapid and efficient killing of *N. fowleri*. Our findings underscore the indispensable role of complement-mediated lysis in host defense against this highly virulent pathogen.

**Figure 4.**
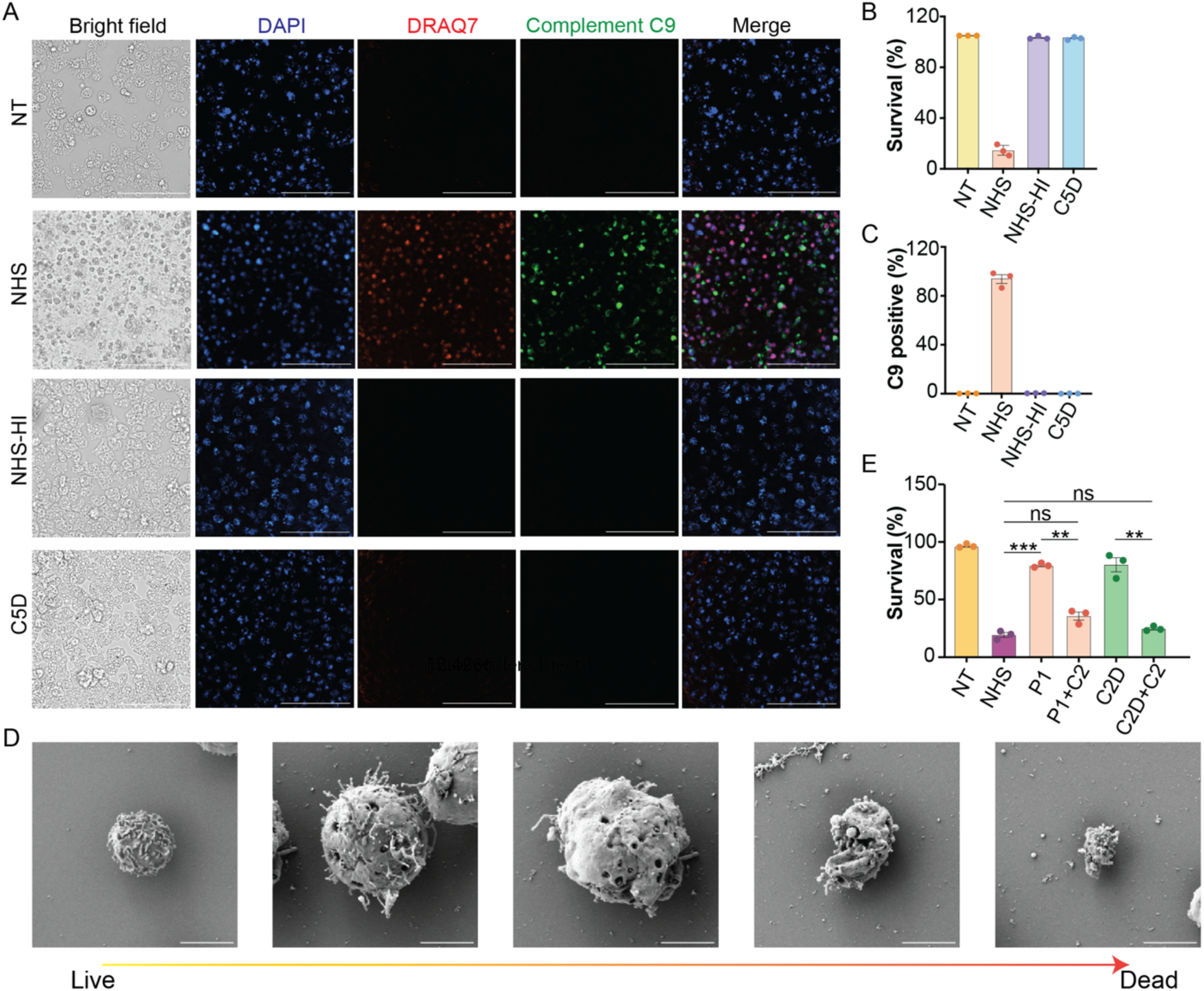
Complement C2 Deficiency Impairs Amoebicidal Activity via Disrupted Classical Pathway Activation. (A) Immunofluorescence microscopy of *N. fowleri* stained for C9 (green), DRAQ7 (red, dead), and DAPI (blue, nuclei) when non-treated (NT) or after treatment with normal human serum (NHS), heat-inactivated normal human serum (NHS-HI), or C5-deficient serum (C5D). Scale bar: 100 µm. (B–C) Quantification of cell death and C9 deposition from (A). (D) Scanning electron microscopy images of *N. fowleri* after normal human serum treatment. Scale bar: 5 μm. (E) Amoebicidal activity of NHS, C2-deficient PAM patient alone or supplemented with purified C2 and serum from C2-deficient patients alone or supplemented with purified C2. Data represent three independent experiments; mean ± SEM. ns: not significant. **P < 0.01; ***P < 0.001 (unpaired t tests). NT: non-treatment; NHS: normal human serum; NHS-HI: normal human serum heat inactivation; C5D: C5 depleted serum

### C2 deficiency leads to impaired amoebicidal activity

Having established the amoebicidal activity of the human complement system, we next sought to determine whether the patient’s serum retained this function. Using *in vitro* viability assays, we observed that the patient’s serum, unlike normal human serum, failed to induce significant killing of *N. fowleri* trophozoites, with survival rates comparable to those in non-treated controls (Figure 4E, S4). To confirm that this defect was specifically attributable to C2, we extended our analysis to sera from other C2-deficient patients (but no clinical history of PAM). These sera similarly lacked amoebicidal activity (Figure 4E, S4), demonstrating that loss of C2 function abolishes complement-mediated lysis of *N. fowleri*. To further confirm the critical and non-redundant role of C2 in anti-*N. fowleri* immunity, we added exogenous purified human C2 protein back to the patient’s serum at increasing concentrations, then incubated it with *N. fowleri* trophozoites. C2 reconstitution restored amoebicidal activity in a dose-dependent manner, with higher C2 concentrations resulting in progressively greater trophozoite killing (Figure S4). At physiological concentration of C2 supplementation, the restored killing efficiency approached that of normal human serum, indicating that C2 alone is sufficient to rescue defective complement-mediated amoebicidal activity in the patient’s serum. Collectively, these data identify C2 as a critical mediator of complement-dependent immunity against *N. fowleri* and establish a direct mechanistic link between genetic C2 deficiency and susceptibility to PAM.

## Discussion

In the past decades, growing evidence has shown that interindividual variability during the course of primary infection can be driven by a monogenic IEI rendering a host permissive to infection.^38,39^ Paradigmatic examples include defects in the TLR3–IRF7 axis predisposing to life-threatening influenza pneumonitis, or deficiencies in the IFN-γ/IL-12 pathway causing Mendelian susceptibility to mycobacterial disease.^40,41^ Despite advances in this field, most severe infectious diseases have not been studied through the lens of IEI, hindering our understanding of the critical and non-redundant players in immunity against these pathogens. Importantly, delineation of these host genetic factors has directly translated into actionable and personalized therapeutic strategies, including interferon supplementation, targeted cytokine blockade, and hematopoietic stem-cell transplantation, with life-saving clinical impact.^42,43^ However, such genotype–phenotype correlations and mechanistic insights remain largely unexplored for most infectious diseases. This knowledge gap is particularly evident in parasitic diseases, which collectively cause millions of deaths worldwide each year.^44^ Progress in understanding parasite pathophysiology has been limited by the intrinsic difficulty of studying these organisms, which often possess complex, multistage life cycles and require distinct intermediate and definitive hosts, complicating experimental modeling and mechanistic dissection.^45–47^ With a mortality rate of approximately 99% when untreated and over 95% when treated, PAM is one of the most lethal infectious diseases known to humans, with very few survivors reported, adding another level of difficulty when it comes to understanding what predisposes individuals to this devastating disease.^48^ Our study of this unique patient transforms our understanding of PAM as we describe for the first time that this devastating parasitic disease can be caused by a monogenic IEI, rather than a random outcome of environmental exposure or generalized immunodeficiency. In light of our findings, considering PAM (and other parasitic diseases) as monogenic diseases could enable genetic diagnosis and personalized treatments for patients, and identify individuals at risk who could implement lifestyle changes to minimize exposure to *N. fowleri*.

The role of the complement system in immunity against *N. fowleri* has remained controversial, with some reports suggesting it plays a role and others suggesting it is redundant.^49,50^ We provide two levels of evidence supporting its critical role in anti-*Naegleria* immunity. At the genetic level, as shown in other infectious diseases, the probability of co-occurrence of a rare genetic disease and an ultra-rare, severe infectious disease without the former causing the latter is extremely low, indicating that C2 deficiency is responsible for PAM in our patient. At the molecular and immunological levels, we have shown that the human complement system can directly kill this parasite and that C2 is critical in this process. Further evidence of this mechanistic link comes from classic mouse studies of PAM in which the A/HeCr mouse strain, which carries a hereditary deficiency in complement component C5, was shown to be the most susceptible to *N. fowleri* infection after intranasal inoculation; mice with depleted complement (e.g., via cobra venom factor) similarly exhibited increased susceptibility, underscoring the protective role of complement in limiting disease *in vivo.*^51^ Complete complement deficiencies were first described in the 1960s and are characterized by the absence or dysfunction of specific complement components, leading to a range of clinical manifestations depending on the affected molecule.^52^ Deficiencies in early components of the classical pathway (such as C1q, C2, and C4) often increase susceptibility to autoimmune diseases, such as systemic lupus erythematosus (SLE). In contrast, deficiencies in terminal components (C5–C9) are strongly associated with recurrent infections by encapsulated bacteria, especially *Neisseria* species.^52,53^ Among complement deficiencies, C2 deficiency is the most common, occurring in roughly 1 in 10,000 to 1 in 20,000 individuals, and representing 29% of all identified complement defects in Western populations. It typically presents with recurrent bacterial infections and autoinflammatory manifestations.^54,55^ We showed that C2 deficiency prevents MAC deposition and effective trophozoite lysis, thereby rendering the host highly vulnerable to infection. In addition, the presence of complement components in CSF, brain tissue, nasal mucus, and epithelial cells suggests that C2 deficiency impairs host defense both at the primary site of infection, the nasal mucosa, and within the central nervous system.^56–59^ Furthermore, the data raise the possibility that other complement component deficiencies might similarly predispose individuals to PAM, expanding the landscape of genetic risk factors for this rare but devastating disease. This conceptual advance reframes PAM as a complement-driven immune defect, opening avenues for improved genetic screening and targeted interventions. By highlighting the indispensable role of complement pathways in controlling *N. fowleri*, our findings provide a mechanistic link between innate immune deficiencies and susceptibility to free-living amoebae, with broad implications for understanding host-pathogen interactions.

PAM remains a nearly uniformly fatal infection, with only 11 documented survivors among 488 reported cases worldwide and only four survivors documented among 167 reported cases in the United States as of 2025.^60,61^ We report an additional survivor with an unusual clinical presentation. Unlike the classical fulminant course of PAM, which is typically fatal within days, and in contrast to nearly all documented survivors who were diagnosed and treated promptly after symptom onset,^62^ this patient survived for over 2 months with ongoing *N. fowleri* infection before initiation of therapy with miltefosine which has been shown to directly kill *N. fowleri* and represent the recommended treatment by the CDC.^60^ The most likely explanation for such an extended disease course is that she was infected with a low-pathogenic amoeba strain. Despite the virulence of different *N. fowleri* strains in humans remain elusive due to the rarity of the disease. Studies in mouse models and *in vitro* characterization of different strains show strain-dependent differences in cytotoxicity and pathogenicity.^63–66^ Hence, infection with a less neurotoxic strain may also have contributed to the indolent disease course and partial neurologic preservation observed in this patient. Our data shows the critical role of the complement system in preventing *N. fowleri* infection. However, the complement system in the context of PAM may work as a double-edged sword. Our findings indicate that the absence of proper complement activation disrupts effective early immune responses, permitting uncontrolled CNS invasion by the pathogen and contributing to disease progression. The importance of complement in the early stages of infection contrasts with its potential to be detrimental once the infection has been established. In fact, part of the brain damage observed in patients with PAM is caused by an exacerbated immune response.^9,67^ Considering the evidence of complement activation and deposition observed in the brain of this PAM patient, and the well-established role of complement in driving neuroinflammation and blood-brain barrier disruption in various central nervous system diseases,^68–71^ we speculate that the complement system antagonistic functions in PAM pathology—contributing both to host defense against *N. fowleri* and to potentially exacerbating neural tissue damage through inflammatory processes. Complement activation in the brain has been implicated in diseases such as Alzheimer’s, multiple sclerosis, and bacterial meningitis, where excessive or dysregulated activation promotes neuronal injury and inflammation,^71^ suggesting a potential similar detrimental role may occur in PAM.

## Conclusions

Primary amoebic meningoencephalitis (PAM) can be caused by an inborn error of immunity, highlighting the importance of genetic factors in its pathogenesis. This case identifies C2 deficiency as a monogenic cause of susceptibility to PAM, establishing the complement system as a critical mediator of human immunity against *N. fowleri*. These findings support integrating complement and genomic analyses into the diagnostic evaluation of PAM.

## Acknowledgments

We would like to thank the patient and her family for taking part in our study. We would also like to thank Dr. Scott Drutman for his help in conceptualizing this study and Dr. Evan Krystofiak for his assistance with the scanning electron microscopy experiments, which were performed in part using the Vanderbilt Cell Imaging Shared Resource (supported by NIH grants CA68485, DK20593, DK58404, DK59637, EY08126, S10OD028704).

## Funding

RMB was funded in part by the National Institute of Allergy and Infectious Diseases (NIAID)(R01AI168210) and the National Cancer Institute (R01CA269217), both part of the National Institute of Health (NIH). CHS was supported by the NIAID (R011AI180777) DER was supported by the NIH T32 GM065086. LIGG was supported by the Instituto de Salud Carlos III (ISCIII) through project FIS-PI21/01642, co-funded by the European Union. LMA and LIGG were supported by the Instituto de Salud Carlos III (ISCIII) through project FIS-PI16/02053, co-funded by the European Union.

## Data availability

The data generated during this study are available from the corresponding author upon reasonable request.

## Author Contributions

Conceptualization: JC, JGM, SRdC, LIG-G, RM-B

Data creation: JC, CR, ND-P, BL, PZ, LMA, CHS, LIG-G, RM-B, JL-M, JMRM, FdFC

Formal analysis: JC, CR

Funding acquisition: LIG-G, RM-B

Investigation: JC, CR, PNA, MD, AVdR, CS, DER

Methodology: JC

Project administration: LIG-G, RM-B

Resources: ND-P, BL, PZ, AVdR, SRdC, ML-T, CHS, RPdD

Supervision: JGM, ML-T, RPdD, CHS, MXB, LIG-G, RM-B

Validation: JC, CR

Visualization: JC, CR, RM-B

Writing original draft: JC, LIG-G, RM-B

All authors reviewed and approved the final draft

## Ethics declaration

## Competing interests

The authors declare no competing interests

## Ethical approval

The study was reviewed and approved by the Vanderbilt University Medical Center Human Research Protection Program (Institutional Review Board # 200412). All participants provided informed consent. All methods were performed in accordance with the ethical standards outlined in the Declaration of Helsinki.

All authors have seen and approved the manuscript

## Notes

### Competing Interest Statement

The authors have declared no competing interest.

