## Supplementary material for "Primary Amoebic Meningoencephalitis caused by Complement C2 Deficiency"

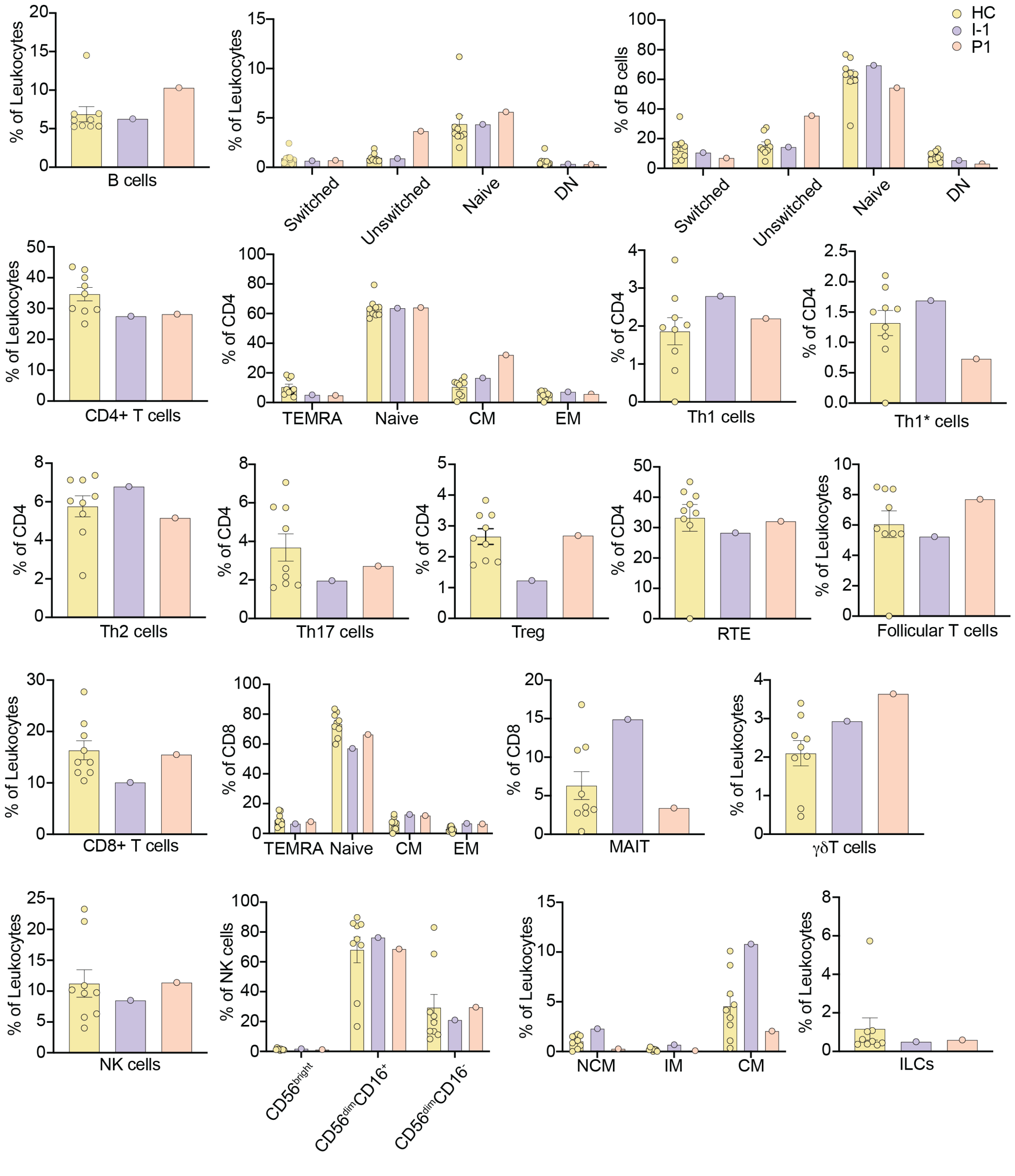


**Supplementary Figure S1. Immunophenotyping frequencies by manual gating.** Frequencies of immune populations as a percentage of leukocytes or parent populations in healthy controls (HC), parent of the patient (I-1), and the patient (P1)

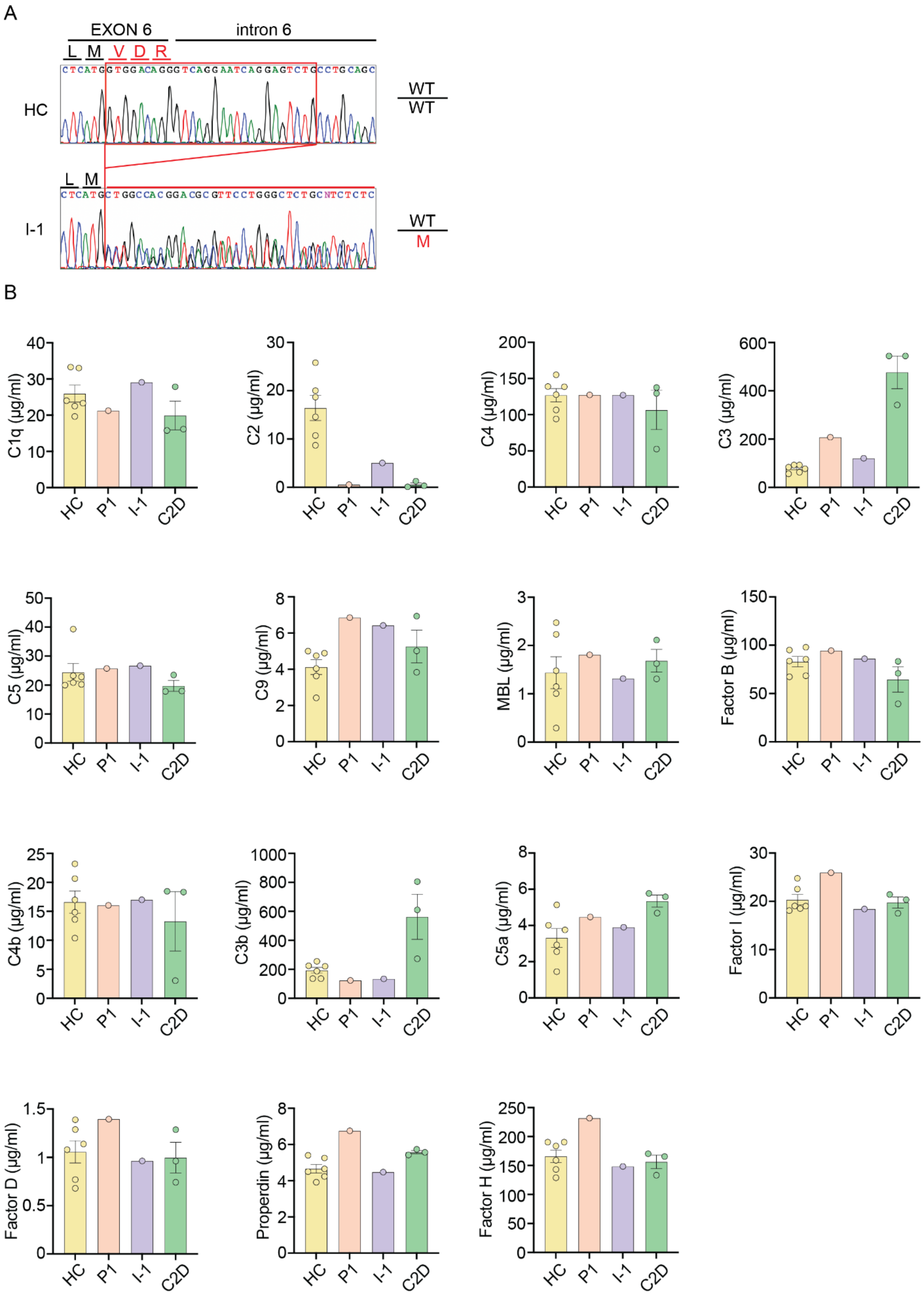
A
EXON 6
intron 6
L M V D R
HC
WT
WT
L M
I-1
WT
M
B
C1q (µg/ml)
C2 (µg/ml)
C4 (µg/ml)
C3 (µg/ml)
C5 (µg/ml)
C9 (µg/ml)
MBL (µg/ml)
Factor B (µg/ml)
C4b (µg/ml)
C3b (µg/ml)
C5a (µg/ml)
Factor I (µg/ml)
Factor D (µg/ml)
Properdin (µg/ml)
Factor H (µg/ml)
HC
P1
I-1
C2D

**Supplementary Figure S2: Genetic characterization and complement component level quantification.**

(A) Sanger sequencing showed a heterozygous mutation in C2 in a parent of the patient (I-1) compared to a healthy control (HC).

(B) Serum levels of complement components and associated factors measured in the patient (P1), a parent of the patient (I-1), and unrelated individuals with C2D, compared to healthy controls (HC).

Each bar represents mean ± SEM; individual data points are shown.

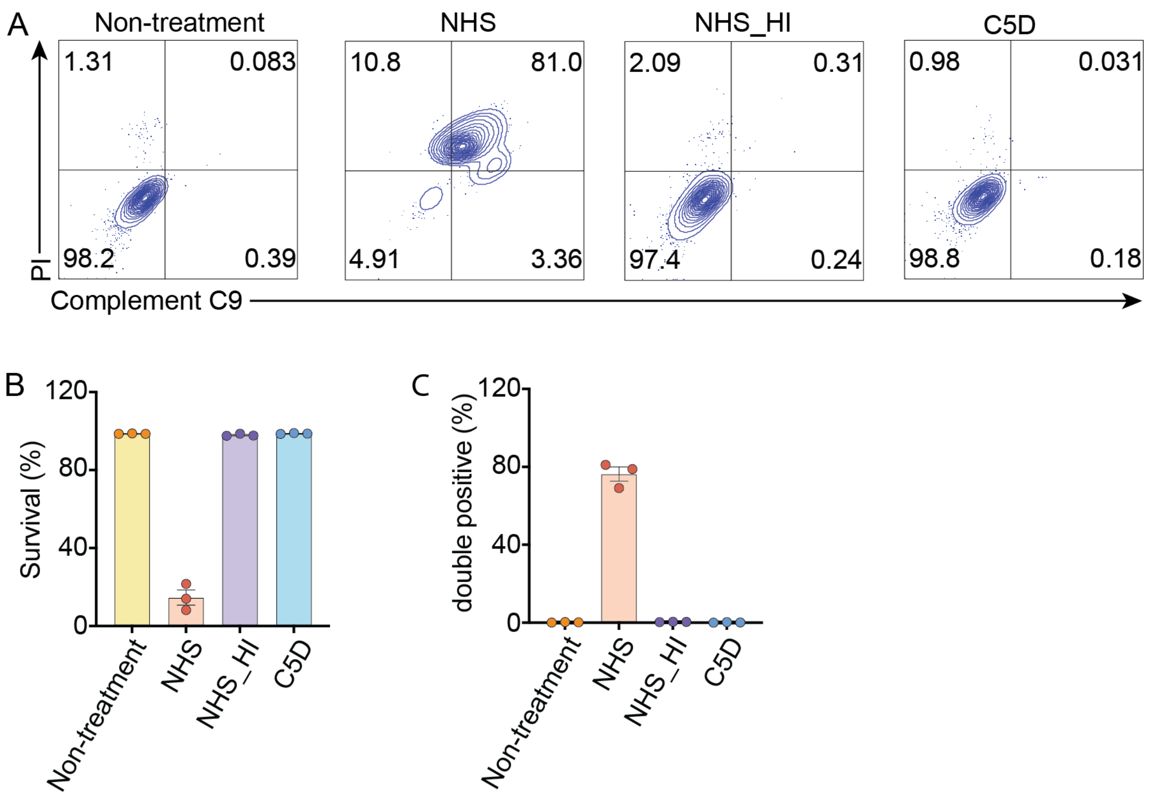


**Supplementary Figure S3: Complement-Mediated Cytotoxicity of *N. fowleri*.**

(A) Flow cytometry analysis of *N. fowleri* trophozoites following treatment with normal human serum (NHS), heat-inactivated NHS (NHS_HI), or C5-depleted serum (C5D). Cells were stained for complement C9 deposition (x-axis) and necrotic cell death using PI (y-axis).

(B) Quantification of trophozoite survival based on exclusion of PI staining.

(C) Percentage of double-positive cells (C9+ PI+) indicating complement-mediated lysis.

Data represent three independent experiments, mean ± SEM from biological replicates.

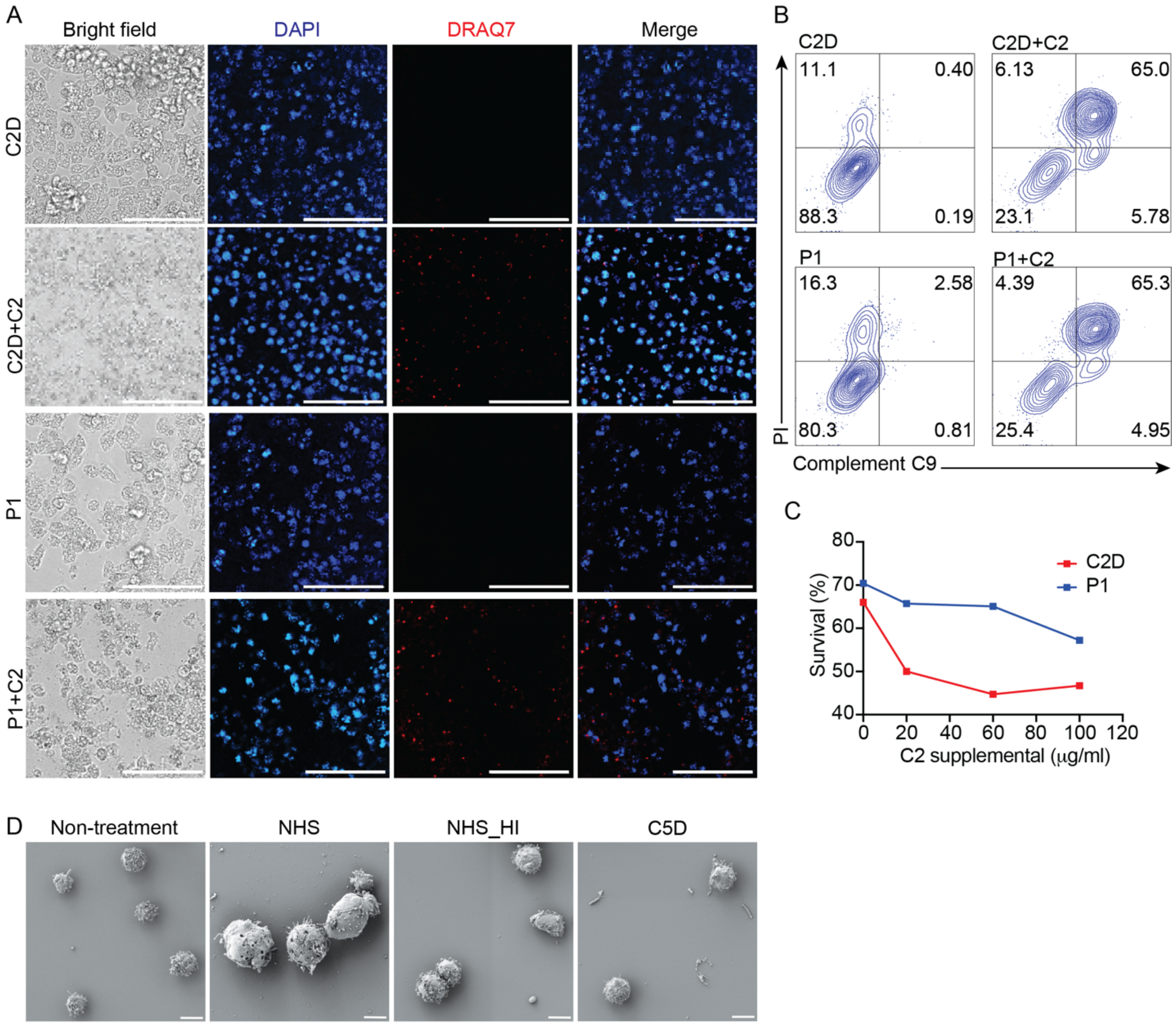


**Supplementary Figure S4: C2 Supplementation Restores Complement-Mediated Killing of *N. fowleri* by C2-Deficient Serum.**

(A) Immunofluorescence microscopy of *N. fowleri* incubated with C2-depleted serum (C2D), serum from the C2-deficient patient (P1), or the same sera supplemented with purified human C2 (C2D+C2, P1+C2). Cells were stained with DAPI (blue, nuclei) and DRAQ7 (red, necrotic/dead cells). Scale bar: 100 μm.

(B) Flow cytometric analysis of complement C9 deposition (x-axis) and cell death (PI, y-axis) on *N. fowleri* treated with the same serum conditions as in A.

(C) Dose-response curve for survival of *N. fowleri* in C2-deficient sera supplemented with increasing concentrations of purified C2.

(D) Electron microscopy of *N. fowleri* with non-treatment or treated with NHS, NHS-HI, and C5D. Scale bar: 10 μm. NHS: normal human serum; NHS-HI: normal human serum heat inactivation; C5D: C5 depleted serum

Data represent three independent experiments, mean ± SEM from biological replicates.

**Supplementary Table S1: Antibodies used for CyTOF**

| Antibody | Clone | Isotype | Dilution | Company |
|---|---|---|---|---|
| Anti-Human CD45 | H130 | 89Y | 1:50 | Standard BioTools |
| Anti-Human CD19 | HIB19 | 142Nd | 1:50 | Standard BioTools |
| Anti-Human CD127/IL7Ra | A019D5 | 143Nd | 1:50 | Standard BioTools |
| Anti-Human CD38 | HIT2 | 144Nd | 1:50 | Standard BioTools |
| Anti-Human IgD | IA6-2 | 146Nd | 1:50 | Standard BioTools |
| Anti-Human CD11c | Bu15 | 147Sm | 1:50 | Standard BioTools |
| Anti-Human CD16 | 3G8 | 148Nd | 1:50 | Standard BioTools |
| Anti-Human CD194/CCR4 | L291H4 | 149Sm | 1:50 | Standard BioTools |
| Anti-Human CD123/IL-3R | 6H6 | 151Eu | 1:50 | Standard BioTools |
| Anti-Human TCRγδ | 11F2 | 152Sm | 1:50 | Standard BioTools |
| Anti-Human CD185/CXCR5 | RF8B2 | 153Eu | 1:50 | Standard BioTools |
| Anti-Human CD3 | UCHT1 | 154Sm | 1:50 | Standard BioTools |
| Anti-Human CD45RA | HI100 | 155Gd | 1:50 | Standard BioTools |
| Anti-Human CD27 | L128 | 158Gd | 1:50 | Standard BioTools |

| | | | | |
|---|---|---|---|---|
| Anti-Human CD28 | CD28.2 | 160Gd | 1:50 | Standard BioTools |
| Anti-Human CD66b | 80H3 | 162Dy | 1:50 | Standard BioTools |
| Anti-Human CD183/CXCR3 | G025H7 | 163Dy | 1:50 | Standard BioTools |
| Anti-Human CD161 | HP-3G10 | 164Dy | 1:50 | Standard BioTools |
| Anti-Human CD45RO | UCHL1 | 165Ho | 1:50 | Standard BioTools |
| Anti-Human CD24 | ML5 | 166Er | 1:50 | Standard BioTools |
| Anti-Human CD197/CCR7 | G043H7 | 167Er | 1:50 | Standard BioTools |
| Anti-Human CD8 | SK1 | 168Er | 1:50 | Standard BioTools |
| Anti-Human CD25 | 2A3 | 169Tm | 1:50 | Standard BioTools |
| Anti-Human CD20 | 2H7 | 171Yb | 1:50 | Standard BioTools |
| Anti-Human HLA-DR | L243 | 173Yb | 1:50 | Standard BioTools |
| Anti-Human CD4 | SK3 | 174Yb | 1:50 | Standard BioTools |
| Anti-Human CD56 | NCAM16.2 | 176Yb | 1:50 | Standard BioTools |
| Anti-Human CD31 | WM59 | 145Nd | 1:50 | Standard BioTools |
| Anti-Human CD196 | G034E3 | 141Pr | 1:25 | Standard BioTools |

| | | | | |
|---|---|---|---|---|
| Anti-Human CD14 | M5E2 | 175Lu | 1:25 | Standard BioTools |
| Anti-Human CD117 | 104D2 | 150Nd | 1:25 | Biolegend |
| Anti-Human TCR Vα24/Jα18 | 6B11 | 156Gb | 1:25 | Biolegend |
| Anti-Human TCR Vα7.2 | 3C10 | 159Tb | 1:25 | Biolegend |
| Anti-Human CD294 | BM16 | 161Dy | 1:25 | Biolegend |

**Supplementary Table S2: primers used for Sanger sequencing**

| | |
|---|---|
| Forward primer | 5' – AAA GCC TGG GCC GTA AAA TCC – 3' |
| Reverse primer | 5' – GAA GAC TTC TTG GAG GAG GTG GG – 3' |

**Supplementary Table S3: primers used for *N. fowleri* PCR test**

| | |
|---|---|
| Forward primer (NFITSFW) | 5’ – TGA AAA CCT TTT TTC CAT TTA CA – 3’ |
| Reverse primer (NFITSRV) | 5’ – AAT AAA AGA TTG ACC ATT TGA AA – 3’ |
| Forward primer (JITSFW) | 5’ – GTC TTC GTA GGT GAA CCT GC – 3’ |
| Reverse primer (JITSRV) | 5’ – CCG CTT ACT GAT ATG CTT AA – 3’ |
